# Pneumococcal Vaccination Coverage and Uptake Among Adults in Switzerland: A Nationwide Cross-Sectional Study of Vaccination Records

**DOI:** 10.1101/2021.10.29.21265674

**Authors:** Kyra D. Zens, Vasiliki Baroutsou, Jan S. Fehr, Phung Lang

**Author notes:** **Correspondence:** Phung Lang.

## Abstract

*Streptococcus pneumoniae*, or pneumococcus, is a common, opportunistic pathogen which can cause severe disease, particularly in adults 65+. In Switzerland, vaccination is recommended for children under 5 and for adults with health predispositions; vaccination of healthy adults 65+ is not recommended. In 2020 we conducted a nationwide, cross-sectional survey of vaccination records to evaluate pneumococcal vaccination coverage and factors affecting uptake among adults 18-85. We found that nationwide coverage was 4.5% without significant regional differences. Coverage was comparable between men and women and between those aged 18-39 (3.0%) and 40-64 (3.2%). Coverage was significantly higher among those 65-85 (9.6%). While 2.7% of individuals reporting no health predisposition were vaccinated, 14.8% with asthma or chronic pulmonary disease, 27.1% with immunosuppression, 12.9% with diabetes, 11.6% with heart, liver, or kidney disease, and 25.9% with >1 health risk were vaccinated. Adjusted odds of vaccination for all health predispositions except heart, liver, or kidney disease were significantly increased. Among unvaccinated individuals “not enough information about the topic” and “not suggested by a doctor/healthcare provider” were the major reasons for abstaining from vaccination. Respondents reporting a health predisposition were significantly less likely to report “not at increased risk due to chronic health conditions or age” as a reason for not being vaccinated (3.7% versus 29.1%) and were more likely to report willingness to be vaccinated in the future compared to those not-at-risk (54.2% versus 39.9%). Our results indicate that pneumococcal vaccination coverage in Switzerland is low among both individuals 65-85 and among those with predisposing health risks. It appears that at-risk individuals are aware of their increased risk, but feel they do not have enough information on the topic to seek vaccination, or have not been recommended a vaccination by their physician.

## 1 Introduction

*Streptococcus pneumoniae*, or pneumococcus, is a common commensal bacterium and opportunistic pathogen. While carriage is typically asymptomatic, pneumococcus can cause a variety of infections including otitis media, sinusitis, and pneumonia [1]. Invasive pneumococcal disease (IPD) occurs when pneumococcus enters normally sterile tissue sites, such as the bloodstream or cerebrospinal fluid, leading to septicemia or meningitis, and can result in significant illness and death [1; 2]. Rates of IPD are highest in children under 5, adults over 65 and individuals with certain chronic health conditions including chronic pulmonary disease, immunosuppression, heart, liver, or kidney disease, or diabetes [3; 4; 5; 6]. In Switzerland, approximately 80% of fatal pneumococcal infections occur in adults 65 and older [7; 8].

Over 90 pneumococcal serotypes have been described. Currently, two types of vaccine are available which protect against a subset of those that most commonly cause disease; a 23-valent polysaccharide vaccine (PPSV23) and the 7- and 13-valent protein-polysaccharide conjugate vaccines (PCV7 and 13) [9]. In Switzerland, both vaccine types are available. Vaccination with PCV13 is recommended for all children under 5 and officially, but “off label” for older children and adults with certain health conditions which predispose for invasive disease including chronic pulmonary disease, immunodeficiency, diabetes, and heart, liver or kidney disease [10; 11] (see Supplementary Figure 1). While PPSV23 was recommended in Switzerland for adults 65+ beginning in 2000, this was rescinded in 2014 due to concerns over efficacy and PPSV23 is no longer officially recommended for any indication [12]. There is currently no recommendation in place for pneumococcal vaccination of otherwise healthy adults, in contrast to the US, the UK and most other EU/EEA countries [13; 14; 15]. These recommendations are, however, highly heterogeneous with regard to both age and risk groups immunized and have further been subject to frequent modifications over the last decade as conjugate vaccines have become available [16; 17].

With an annual incidence of approximately 10 cases per 100,000 individuals for IPD alone [18], pneumococcus remains an important cause of vaccine-preventable infections in Switzerland. For comparison, in a recent study of European surveillance data assembled from 29 countries, crude IPD incidence in 2018 was 6.4 cases per 100,000 (range 0.2 to 16 cases per 100,000) [6]. According to the Swiss National Vaccination Coverage Survey, which evaluates vaccination coverage in children and adolescents, 84% of 2-year-olds and 75% of 8-year-olds had received at least one PCV dose between 2017 and 2019 [19]. There is, however, no equivalent vaccination monitoring system in Switzerland for adults and relatively little is known regarding adult pneumococcal vaccine coverage. Whether at-risk individuals are appropriately vaccinated is not known and how health risk factors predisposing for pneumococcal infection impact pneumococcal vaccine uptake is unclear. Here, we conducted a nationwide, cross-sectional study to assess pneumococcal vaccination and factors affecting uptake among adults in Switzerland.

## 2 Methods

### 2.1 Selection and Recruitment of Participants

We conducted a nationwide, cross-sectional study based on obtaining vaccination records by mail [19; 20]. In brief, adults between 18 and 85 with a Swiss mailing address were divided into 3 age groups (18-39, 40-64, 65-85) and selected from each of the 7 Swiss geographical “large regions” (as defined by the Swiss Federal Statistical Office [21], Supplementary Table 1) by disproportionate stratified random sampling. From each age group/region, 1,280 individuals were invited to participate (n=26,880). Potential participants were contacted by mail beginning in March 2020. To participate, individuals were requested to submit a copy of their vaccination record and to complete a short questionnaire. Non-responders received a single reminder letter in August 2020. As Switzerland is a multilingual country, the language (German, French, or Italian) of both the letter and questionnaire were matched to the official language of the community of each address (Supplementary Table 2). Communities in which the official language was Romansch (0.3% of population) were excluded. The total enrollment period was from March to December 2020. Participants submitting a completed questionnaire, vaccination record, or both, were eligible for a lottery to receive one of three 1,000 CHF ($1,065) prizes with individuals submitting both entered twice.

### 2.2 Data Collection

#### 2.2.1 Vaccination Records

Both paper-based forms and private digital vaccination records (i.e. from a healthcare provider) are used in Switzerland. Vaccination records contain dates of vaccination, vaccine names and lot numbers that are manually recorded by a health professional after a vaccination is administered. For this study, participants could submit a copy of their vaccination record and completed questionnaire by mail or online by email or via our digital platform. Copies of paper-based forms or printouts of digital vaccination records obtained from study participants were manually inspected and the number, date(s), and formulation(s) of pneumococcal immunization were recorded.

#### 2.2.2 Self-Report Questionnaire

Participants were asked to report 1) Whether they had experienced one of the following health risks: asthma or a chronic respiratory condition; immunosuppression due to a medical condition or treatment; heart, liver, or kidney disease; diabetes; 2) Whether they smoke >1 cigarette or vape >1mL a day; 3) Whether they have one of the following occupational exposures: work in a healthcare setting with frequent patient contact; or work with children under the age of 5; and 4) If they have not been vaccinated for pneumococcus, what had prevented them and whether they would consider doing so in the future. Completed questionnaires were manually inspected and responses recorded.

### 2.3 Ethical Considerations

With each mailing, a letter explaining the study’s procedures and objectives was included. In this letter, individuals were informed that study participation was voluntary and that they had the possibility to withdraw submitted data at any time. They were informed that, by submitting completed questionnaires and/or vaccination records, they were consenting to participation in the study. All data were treated confidentially and anonymized prior to analysis. The study procedure and method of consent were approved by the Department of Data Protection of the University of Zurich and the Ethics Committee of the Canton of Zurich.

### 2.4 Statistical Analyses

Prior to analysis, data were adjusted for study design and non-response and post-stratified by age and gender, such that they were representative of the population of Switzerland. 2019 population data for age and gender by region used to calculate analysis and post-stratification weights were obtained from the Swiss Federal Statistical Office [22]. Vaccination coverage among study participants was calculated by dose counting. As pneumococcal vaccination for adults is, in nearly all instances, recommended as a single dose (both currently for PCVs in “at-risk” individuals [13] and previously for PPSV23 for adults 65+ [23]), we considered receipt of one or more pneumococcal vaccine doses as “vaccinated”, Descriptive statistics with 95% confidence intervals were calculated for vaccination coverage for both demographic and self-reported risk variables from the questionnaire. Vaccination coverage by linguistic or geographical region was evaluated by linear regression using the Adjusted Wald test and Bonferroni’s adjustment. A multivariable logistic regression model was constructed to evaluate the association between age, gender, the presence of self-reported health risks with the probability of being vaccinated and crude and adjusted Odds Ratios (OR) with 95% confidence intervals calculated. To evaluate the timing of vaccination and vaccine formulation usage, vaccination date and formulation data from vaccinated individuals were used. For analysis of reasons for not vaccinating and intention to vaccinate, questionnaire data were evaluated from non-vaccinated individuals and stratified by the presence or absence of self-reported health risks. Frequencies of expected versus observed values were compared by Pearson’s χ^2^ test using the Rao & Scott adjustment. Vaccination coverage by geographic region was visualized using R v.4.0.5 (R Foundation for Statistical Computing, Vienna, Austria). Statistical analyses were performed using Stata v.17.0 (StataCorp, LLC, College Station, TX, USA) and GraphPad Prism v.8.0 (GraphPad Software, Inc., San Diego, CA, USA); p values <0.05 were considered statistically significant.

## 3 Results

### 3.1 Participation

Of 26,880 randomly selected individuals which were contacted, 7,761 (28.9%) responded. Of these, 3,503 submitted a questionnaire without a vaccination record and were excluded from further analysis (Supplementary Figure 2). A total of 4,258 individuals (15.8%, Supplementary Figure 2, Table 1) submitted both a completed questionnaire and vaccination record and were included for analysis. For simplicity, we will refer to these individuals as “participants” or “respondents”. 17.5% of invited women and 14.1% of invited men participated; 55.6% of participants were female (Table 1). Participation by age group was 12.9% for those 18-39, 16.3% for those 40-64 and 18.2% for those 65-85 (Table 1). Participants tended to be older in comparison to the adult population of Switzerland (Table 1). Regionally, participation was highest in Zurich and lowest in Lake Geneva (22.9 and 8.7% of invited individuals, respectively, Table 1).

**Table 1.**
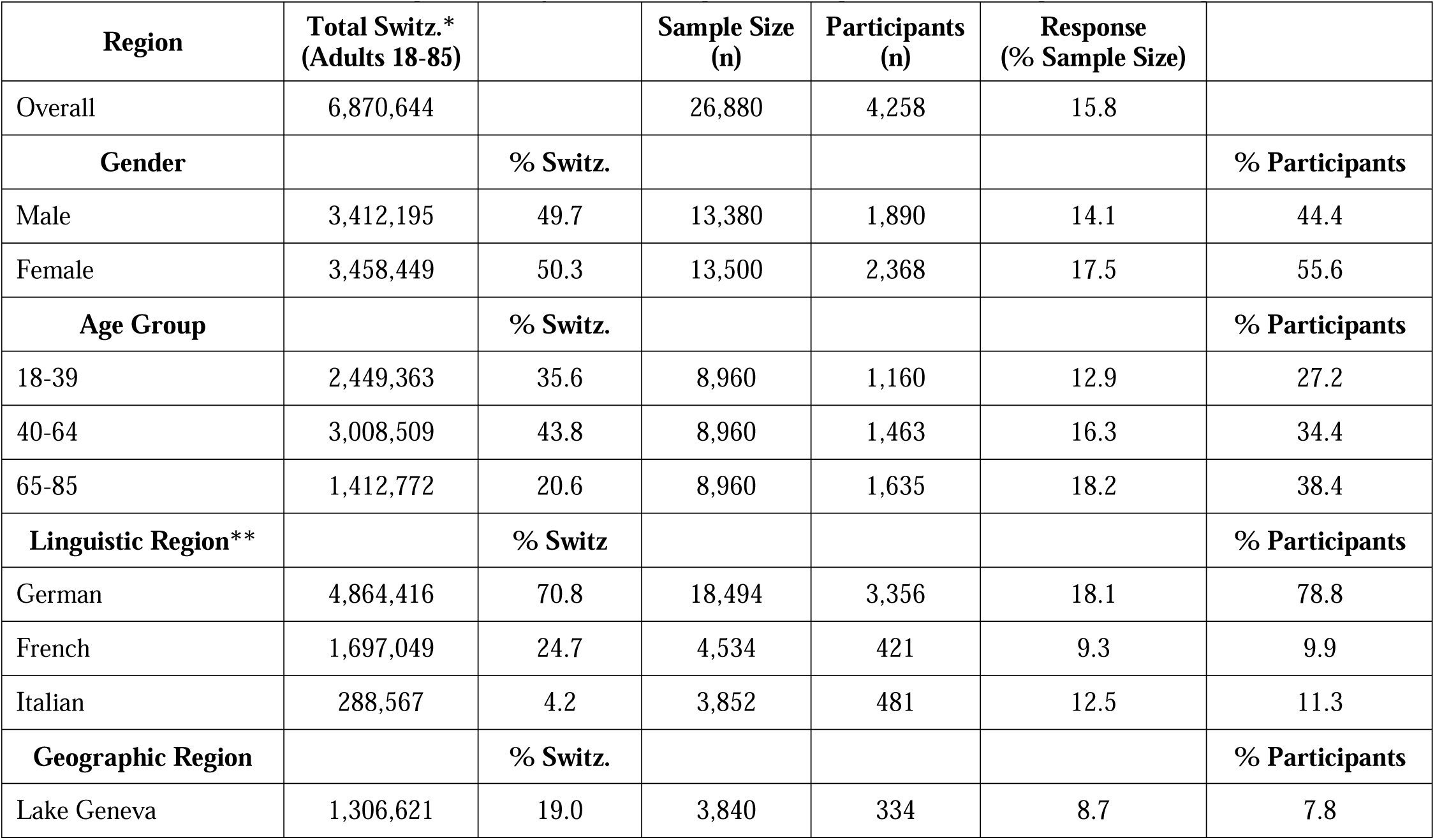
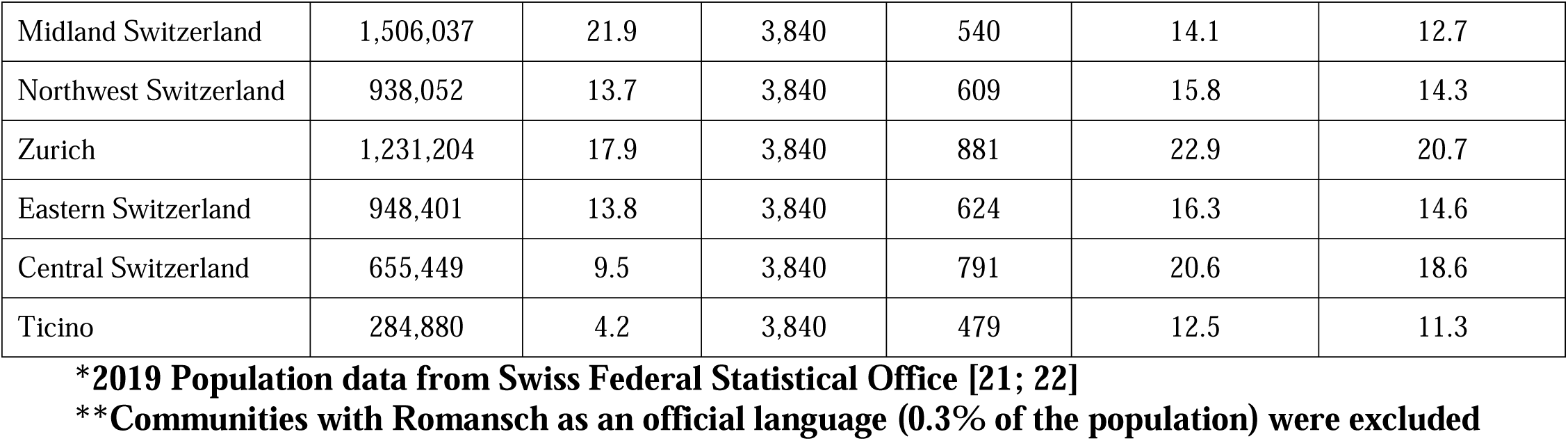
Participant Demographics by Gender, Age and Linguistic and Geographical Region

| Region | Total Switz.*<br>(Adults 18-85) |  | Sample Size<br>(n) | Participants<br>(n) | Response<br>(% Sample Size) |  |
| --- | --- | --- | --- | --- | --- | --- |
| Overall | 6,870,644 |  | 26,880 | 4,258 | 15.8 |  |
| **Gender** |  | **% Switz.** |  |  |  | **% Participants** |
| Male | 3,412,195 | 49.7 | 13,380 | 1,890 | 14.1 | 44.4 |
| Female | 3,458,449 | 50.3 | 13,500 | 2,368 | 17.5 | 55.6 |
| **Age Group** |  | **% Switz.** |  |  |  | **% Participants** |
| 18-39 | 2,449,363 | 35.6 | 8,960 | 1,160 | 12.9 | 27.2 |
| 40-64 | 3,008,509 | 43.8 | 8,960 | 1,463 | 16.3 | 34.4 |
| 65-85 | 1,412,772 | 20.6 | 8,960 | 1,635 | 18.2 | 38.4 |
| **Linguistic Region\*\*** |  | **% Switz** |  |  |  | **% Participants** |
| German | 4,864,416 | 70.8 | 18,494 | 3,356 | 18.1 | 78.8 |
| French | 1,697,049 | 24.7 | 4,534 | 421 | 9.3 | 9.9 |
| Italian | 288,567 | 4.2 | 3,852 | 481 | 12.5 | 11.3 |
| **Geographic Region** |  | **% Switz.** |  |  |  | **% Participants** |
| Lake Geneva | 1,306,621 | 19.0 | 3,840 | 334 | 8.7 | 7.8 |
| Midland Switzerland | 1,506,037 | 21.9 | 3,840 | 540 | 14.1 | 12.7 |
| Northwest Switzerland | 938,052 | 13.7 | 3,840 | 609 | 15.8 | 14.3 |
| Zurich | 1,231,204 | 17.9 | 3,840 | 881 | 22.9 | 20.7 |
| Eastern Switzerland | 948,401 | 13.8 | 3,840 | 624 | 16.3 | 14.6 |
| Central Switzerland | 655,449 | 9.5 | 3,840 | 791 | 20.6 | 18.6 |
| Ticino | 284,880 | 4.2 | 3,840 | 479 | 12.5 | 11.3 |

**\*2019 Population data from Swiss Federal Statistical Office [21; 22]**
**\*\*Communities with Romansch as an official language (0.3% of the population) were excluded**

### 3.2 Vaccination Coverage by Gender, Age Group, and Region

Among participants, nationwide adult pneumococcal vaccination coverage (at least one dose) was 4.5% (95%CI 3.8-5.2, n=4,258). Vaccination coverage was comparable between men and women (4.8% vs 4.2%, p=0.36, Table 2). Coverage was also comparable between those aged 18-39 (3.0%) and 40-64 (3.2%), but was higher among those 65-85 (9.6%, p<0.001, Table 2). Vaccination coverage was similar by linguistic region; 4.5% in German-speaking areas and 4.2% in French and Italian-speaking areas (p=0.83, Table 2). By geographic region, vaccination coverage ranged between 2.5% and 5.8% (Eastern and Midland Switzerland, respectively), though these differences were not significant (p=0.11, Supplementary Figure 3).

**Table 2.** Logistic Regression of Sex, Age Group and Self-Reported Health Risk Factors on Adult Pneumococcal Vaccination Coverage

| **Gender** | **% Vaccinated**<br>**(95%**<br>**Confidence)** | **Crude Odds Ratio**<br>**(95% CI)** | **p Value** | **Adjusted Odds Ratio**<br>**(95% CI)** | **p Value** |
| --- | --- | --- | --- | --- | --- |
| Male | 4.80<br>(3.79-6.07) | 1.17<br>(0.84-1.61) | 0.357 | Ref. | -- |
| Female | 4.15<br>(3.37-5.09) | 0.86<br>(0.62-1.18) | 0.357 | 0.78<br>(0.55-1.11) | 0.164 |
| **Age Group** |  |  |  |  |  |
| 18-39 | 3.04<br>(2.02-4.56) | 0.56<br>(0.36-0.89) | 0.014 | Ref. | -- |
| 40-64 | 3.24<br>(2.35-4.44) | 0.58<br>(0.40-0.85) | 0.005 | 0.88<br>(0.51-1.52) | 0.649 |
| 65-85 | 9.59<br>(8.13-11.13) | 3.26<br>(2.38-4.48) | <0.001 | 2.48<br>(1.53-4.00) | <0.001 |
| **Disease** |  |  |  |  |  |
| No Risk Reported | 2.67<br>(2.09-3.40) | 0.17<br>(0.12-0.24) | <0.001 | Ref. | -- |
| Asthma | 14.75<br>(11.24-19.12) | 4.65<br>(3.21-6.73) | <0.001 | 5.09<br>(3.28-7.91) | <0.001 |
| Immunosuppression | 27.10<br>(18.18-38.33) | 9.01<br>(5.23-15.53) | <0.001 | 16.10<br>(8.20-31.60) | <0.001 |
| Diabetes | 12.86<br>(7.75-20.60) | 3.35<br>(1.86-6.03) | <0.001 | 3.32<br>(1.48-7.45) | 0.004 |
| Heart/Liver/Kidney Disease | 11.53<br>(7.96-16.43) | 3.07<br>(1.96-4.81) | <0.001 | 1.26<br>(0.65-2.44) | 0.500 |
| >1 Health Risk | 25.91<br>(16.65-37.96) | 8.15<br>(4.53-14.67) | <0.001 | 9.69<br>(5.19-18.09) | <0.001 |

### 3.3 Vaccination Coverage by Risk Factors

Using questionnaire data, we evaluated pneumococcal vaccination coverage among individuals self-reporting one or more of health risk factors (n=813, Supplementary Table 3). We found that 14.8% of individuals reporting asthma, 27.1% reporting immunosuppression, 12.9% reporting diabetes, and 11.6% reporting heart, liver, or kidney disease, were vaccinated. 25.9% of individuals reporting more than one health risk were vaccinated compared to 2.7% of those not reporting any health risk (Table 2). Independently, asthma, immunosuppression, diabetes, or heart/liver/kidney disease were associated with 4.7, 9.0, 3.4 and 3.1-fold increased odds of pneumococcal vaccination, respectively (Table 2). Having more than one health risk was associated with 8.2-fold increased adjusted odds of vaccination (Table 2). Adjusted odds of pneumococcal vaccination for all health risks except heart/liver/kidney disease were significantly increased (Table 2). Adjusted odds of pneumococcal vaccination for those with more than one health risk were also significantly increased (Table 2). In evaluating health risk by vaccine type, 9.8% of those vaccinated with PCV7 reported a health risk, compared to 59.7% of those vaccinated with PCV13 and 45.0% of those vaccinated with PPSV23.

### 3.4 Timing of Pneumococcal Vaccination and Vaccine Type

Among vaccinated individuals (n=230) we assessed the year of first pneumococcal vaccination by age to identify possible temporal trends in uptake. Points a and b in Figure 1A indicate the period from 2000-2014 when vaccination was recommended in Switzerland for all individuals 65+. We observed a peak in pneumococcal vaccination in 2009-2010 and a trend toward increased vaccination beginning in 2014, with the highest level in 2020. As our study was conducted in 2020, however, data for this year are not complete.

**Figure 1.**
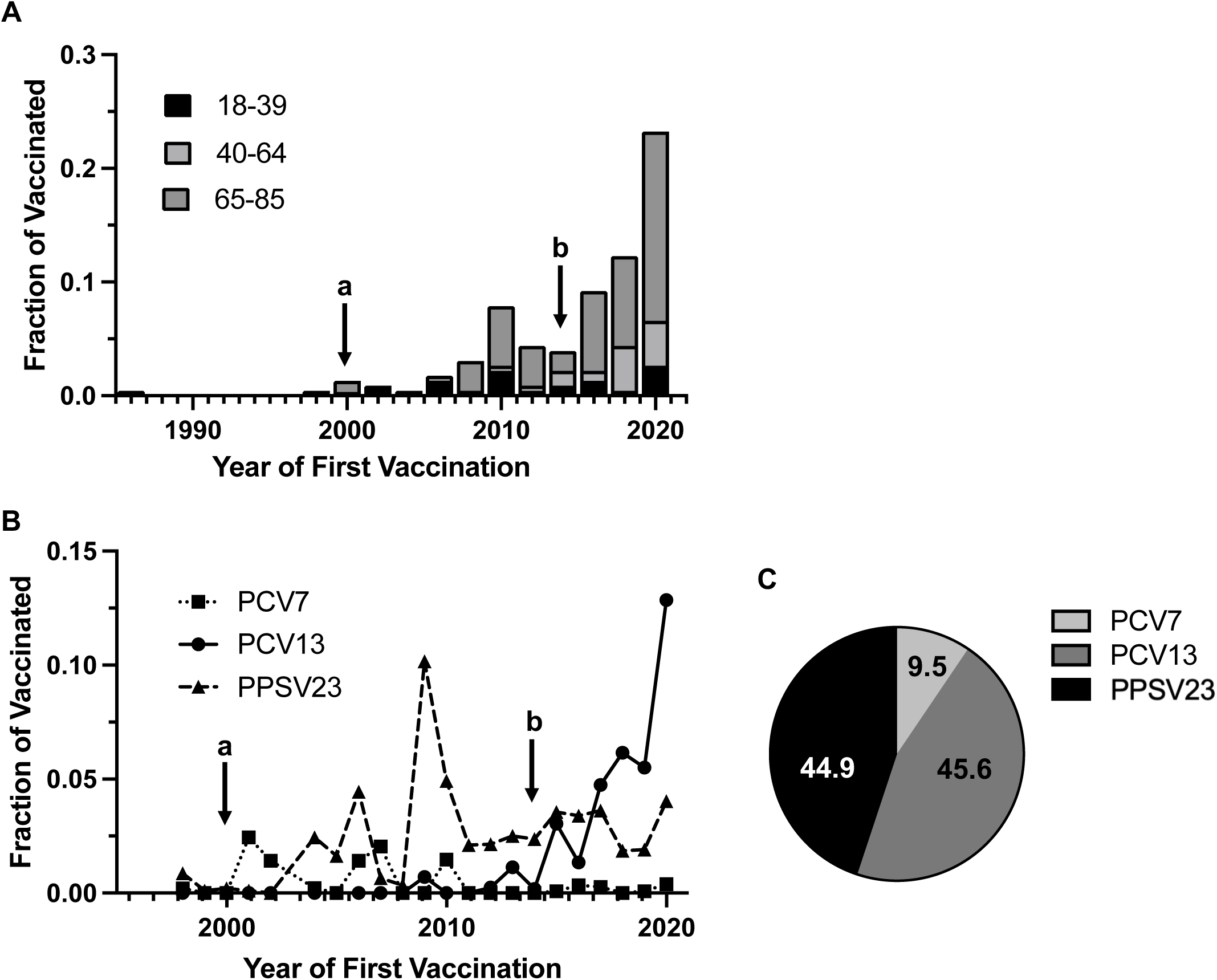
Timing of Pneumococcal Vaccination and Vaccine Type Usage. In Figure 1A and 1B, point (a) denotes official recommendation of PPSV23 for adults 65+ and those at risk of severe disease in Switzerland; point (b) denotes an end to the official recommendation for vaccination of adults 65+ and a switch to PCV13 for those at risk of severe disease, (n=230, 2 values were excluded because of missing date information in the vaccination record). Note that 2020 was a partial study year. **(A)** Year of First Vaccination: The fraction of individuals receiving at least one pneumococcal vaccine dose which received their first pneumococcal vaccination in a given year, by age group. **(B)** Vaccine Type Usage by Year: The fraction of first pneumococcal vaccine doses in a given year, by vaccine type. **(C)** Total Pneumococcal Vaccine Doses by Type: The percentage of total pneumococcal vaccine doses received by individuals, by vaccine type (n=297 doses from 230 individuals, 11 values were excluded because of missing vaccine type information in the vaccination record).

We also evaluated the year and age of first pneumococcal vaccination by vaccine type (Figure 1B). Among study participants PCV7, PCV13, and PPSV23 were used. Of all pneumococcal vaccine doses received by study participants (n=297), 9.5% (95%CI 6.4-13.3) were with PCV7, 45.6% (95%CI 39.7-51.3) were with PCV13, and 44.9% (95%CI 38.4-50.0) were with PPSV23 (Figure 1C). Use of PCV7 was generally low, accounting for a maximum of 2.4% of first doses in 2001. 24.6% (95%CI 5.6-47.5) of first vaccinations with PCV7 took place when individuals were aged 5 or younger. Use of PCV13 increased consistently since 2012, reaching 5.5% and 12.9% of total first doses in 2019 and 2020, respectively. All study participants receiving PCV13 as a first vaccination were first vaccinated after age 5. PPSV23 accounted for 4.5%, 10.2% and 4.9% of first doses in 2006, 2009 and 2010, respectively. From 2011, usage ranged from 1.9% to 3.6%. In 2020 PPSV23 represented 4.0% of total first doses. 4.2% (95%CI 0.1-6.4) of first vaccinations with PPSV23 occurred when individuals were aged 5 or younger.

### 3.5 Vaccine Perception and Intention to Vaccinate

Among unvaccinated individuals (n=4,028) we evaluated reasons for abstaining from pneumococcal vaccination, asking participants to indicate which reason(s) had prevented them from being vaccinated and stratifying on the presence of one or more reported health risk factors (Figure 2A). In general, results were comparable between unvaccinated individuals reporting a health risk and those not reporting such a risk. We found that 70.1% (95%CI 65.8-74.1) of respondents with a health risk and 64.7% (95%CI 62.7-66.6) without listed “not enough information about the topic” as a reason for not receiving pneumococcal vaccination. 44.4% (95%CI 40.0-49.0) of those at-risk and 42.9% (95%CI 40.9-44.8) of those not-at-risk listed “not suggested by a doctor or healthcare provider”.

**Figure 2.**
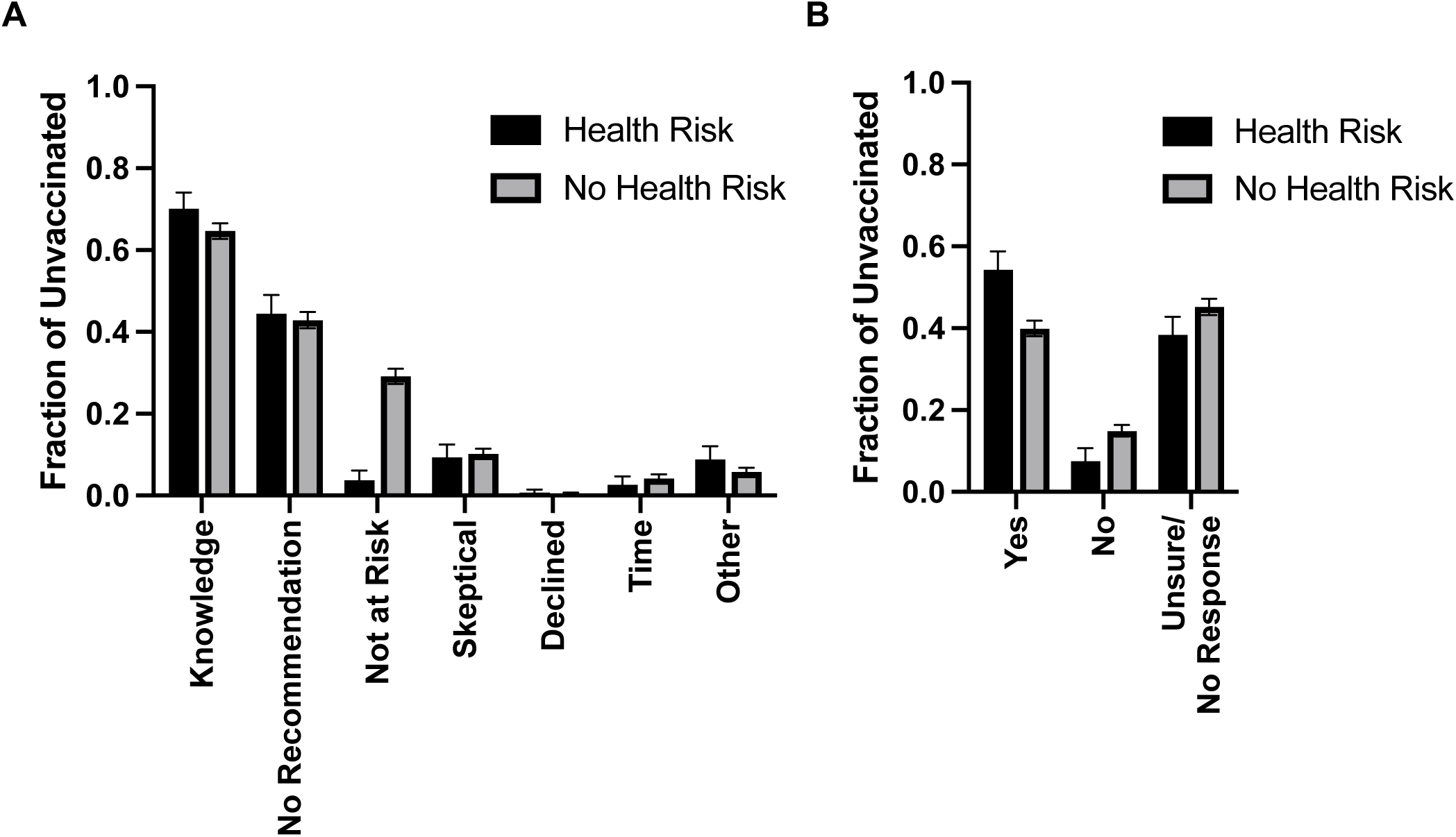
Reasons for Not Vaccinating and Intention to Vaccinate Among Unvaccinated. **(A)** Reasons for not obtaining pneumococcal vaccination among unvaccinated: The fraction of unvaccinated individuals reporting the indicated reasons for not obtaining pneumococcal vaccination, stratified by individuals reporting one or more health risks, or individuals reporting no health risks (n=4,028). **(B)** Willingness to Receive Pneumococcal Vaccination: The fraction of unvaccinated individuals reporting willingness for obtaining pneumococcal vaccination in the future, stratified by individuals reporting one or more health risks, and individuals reporting no health risks (n=4,028).

Similarly, approximately 10%, 0.5%, 4% and 6% of participants with and without health risks listed that they were “skeptical of vaccines”, had “declined vaccination”, had “not enough time or interest”, or “other”, as reasons for not vaccinating (Figure 2A). We did observe, however, that respondents reporting a health risk were significantly less likely to report “not at increased risk due to chronic health conditions or age” as a reason for not being vaccinated compared to those not reporting a health risk (3.7%, 95%CI 2.3-6.2, versus 29.1%, 95%CI 27.3-31.0, p<0.001).

We further investigated the willingness of these unvaccinated individuals to be vaccinated against pneumococcus, asking whether they “would consider receiving the vaccine in the future”. Again, we stratified answers by respondents’ health risk status (Figure 2B). We found that significantly more at-risk unvaccinated individuals reported that they were willing to be vaccinated compared to not-at-risk unvaccinated individuals (54.2%, 95%CI 49.6-58.7, versus 39.9%, 95%CI 38.0-41.9, respectively; p<0.001). Consistent with this, we observed a corresponding decrease in at-risk individuals reporting that they would not receive the vaccine in the future (7.5%, 95%CI 5.3-10.6, versus 14.9%, 95%CI 13.5-16.4 not-at-risk) and those reporting that they were “unsure” or having no response (38.3%, 95%CI 34.0-42.7, at-risk versus 45.2%, 95%CI 43.2-47.2, not-at-risk).

## 4 Discussion

Pneumococcal infections remain a leading cause of morbidity and mortality among adults, particularly in those 65+ or with predisposing health risks. To our knowledge, this is the first study to evaluate pneumococcal vaccination of adults, and factors affecting vaccination uptake, throughout Switzerland.

### 4.1 Vaccination Coverage by Gender, Age Group, and Region

As a result of recent modifications and heterogeneity among national adult pneumococcal vaccination recommendations [16; 17], there is a need for reliable estimates of current coverage in many countries. Here, we found that overall adult pneumococcal vaccination coverage in Switzerland was 4.5% and did not differ significantly by gender or linguistic or geographical region.

Interestingly, in a recent study of pneumococcal vaccination in children, vaccination uptake was slightly higher in “west” (mostly German-speaking) compared to “east” (mostly French-speaking) Switzerland. [24] While coverage was comparable in our study between individuals 18-39 and 40-64 (approximately 3%), it was significantly higher among those 65-85, nearing 10%.

Previous work has demonstrated a clear trend toward increased overall pneumococcal vaccination coverage in countries recommending vaccination for both elderly and at-risk individuals compared to countries recommending vaccination of only those at risk [25]. France, which neighbors Switzerland, recommends pneumococcal vaccination only for at-risk adults. Coverage among healthy adults 18-64 and 65+ was estimated at just under 1%, and just over 3%, respectively [26], while studies among those 65+ in care facilities and at high risk of disease found that coverage in this group was approximately 20% [27; 28; 29]. In contrast, Germany and Italy, which also border Switzerland, recommend pneumococcal vaccination for both at-risk and elderly individuals. In a study of German health insurance claims data, coverage among those 18-59 was just under 4% compared to 51% among those 60+ [30]. Similarly, in Italy, overall PPSV23 coverage was 680 doses per 10,000 individuals (which could be roughly interpreted as approximately 7%) [25], while a study of regional health registry data found that approximately 30% of those 65+ and 26% of those with health risks had been vaccinated [31]. In the United States and United Kingdom, which also recommend vaccination of elderly individuals, coverage among those 65+ has been estimated at 62% and 70%, respectively [32; 33].

As pneumococcal vaccination was recommended in Switzerland for adults 65+ without additional health risk factors from 2000-2014 [12], that we observe increased, but still low, coverage among those 65-85 is, perhaps, unsurprising. Importantly, though, individuals 65+ are among those most at-risk of severe pneumococcal disease [34; 35]. In a study of surveillance data compiled from 29 EU/EEA countries, but not including Switzerland, the overall IPD notification rate among those 65+ was 18.7 per 100,000 with a 21% fatality rate [6]. IPD notification rates among those 65+ in France, Italy and the United Kingdom (Germany does not have a mandatory national reporting system) were 20.9, 6.7 and 28.8 cases per 100,000, respectively [6]. In comparison, IPD incidence in Switzerland among those 65+ was approximately 50 per 100,000, ranging from 21 per 100,000 for those 65-69 and 88 per 100,000 for those 95+ [18]. While it is important to note that reporting differs between countries and these values are not necessarily directly comparable, these findings serve to highlight a need for making effective decisions regarding pneumococcal vaccination in this age group a priority.

### 4.2 Vaccination Coverage by Risk Factors

In evaluating pneumococcal vaccination coverage among individuals reporting one or more health risk(s) predisposing for IPD, we found that chronic respiratory disease, immunosuppression, diabetes, or having more than one of these risks were all associated with increased likelihood of pneumococcal vaccination. These conditions were all found to be independent risk factors for pneumococcal pneumonia in Switzerland, supporting their use as grounds for vaccination [34]. In our study, however, pneumococcal vaccination coverage in these groups was still low; only ∼15% of individuals reporting respiratory disease, ∼27% with immunosuppression, ∼12% with heart, liver, or kidney disease, and ∼13% with diabetes were vaccinated, despite their eligibility. Just 26% of individuals with multiple health risks were vaccinated.

A 2021 study of claims in the main French national health insurance scheme found that approximately 10% of all adults had a risk condition predisposing for pneumococcal disease [36]. However, an analysis published by the French High Council for Public Health estimated that only 20% of at-risk adults are vaccinated [37]. In a separate study of claims data ∼12% of adult HIV patients, ∼7% of chronic obstructive pulmonary disease or congestive heart failure patients, and ∼2% of those with diabetes were vaccinated within 2 years of diagnosis, although vaccination was indicated for these conditions [26]. A similar study in Germany found the overall vaccination rate among newly-diagnosed, at-risk patients was ∼4% within 2 years with the highest rates in patients starting immunosuppressive therapy for rheumatoid arthritis (12%) and in those with HIV (10%) [38]. Vaccination coverage among German adults 18-59 with at least one health risk factor was just under 13% [30]. Among those 60+, however, this increased to nearly 55% [30]. Similarly, 13% of US adults 18-64 with diabetes and 15% of those with lung disease were vaccinated for pneumococcus [39]; coverage was estimated at 20% overall in high-risk patients younger than 65 [40]. Importantly, these findings indicate that vaccination is increased among those at highest risk in Switzerland, but that at-risk individuals remain substantially under-vaccinated, both in Switzerland, and elsewhere.

### 4.3 Timing of Vaccination and Vaccine Type Used

Among vaccinated individuals we observed peaks in receiving first pneumococcal vaccine doses in 2009-2010, which correlates with the 2009-2010 H1N1 influenza pandemic, and in 2020, which correlates with the SARS-CoV-2 pandemic. As pneumococcus is an important cause of secondary bacterial pneumonia following viral respiratory infection [41; 42], a severe viral respiratory disease season could be grounds for vaccination.

Of administered pneumococcal vaccine doses, ∼10% were PCV7, 46% PCV13, and 45% PPSV23. Use of PCV7 was consistently low, ranging between 0 and 2.4% of first doses annually. This is to be expected as, while PCV7 was recommended from 2001 for all children <5 with predisposing health risks and for all children <2 between 2005 and 2010 [43], there was no PCV7 recommendation for adults at any time. Therefore, PCV7 vaccinations observed in our study would be expected to be received in early childhood. Interestingly, while ∼25% of first vaccinations with PCV7 were in individuals when they were aged 5 or younger, the majority of first vaccinations with PCV7 were in older individuals, suggesting “off-label” usage in older children and adults. In 2010 PCV7 was officially replaced by PCV13 and the recommendation for pneumococcal vaccination was extended to all children <5. Use of PCV13 among study participants increased consistently from 2010, exceeding PPSV23 doses in 2017. The recommendation for PCV13 vaccination was extended to adults with predisposing health risks in 2014 [12]. Between 2000 and 2014, PPSV23 was officially recommended for routine vaccination of otherwise healthy adults aged 65+ [12]; its usage peaked in 2009-2010.

Importantly, PPSV23 is based on polysaccharide antigens which elicit a so-called T cell-independent antibody response. Such responses are generally short-lived (typically lasting a few years) and are not “recalled” by booster immunizations. In contrast, polysaccharide-protein conjugate vaccines (PCV7 and 13) produce T cell-dependent antibody responses which tend to be more durable and can be boosted by subsequent immunization. It is generally thought that older individuals and individuals with health conditions compromising immune function tend to not respond robustly to polysaccharide antigens (such as PPSV23), thus limiting their effectiveness. Based in part on this, it was recommended in Switzerland to discontinue general vaccination of those 65+ with PPSV23 and to vaccinate only those adults (aged 18+) with health risks with PCV13 [12].

Whether a general PCV13 vaccination recommendation in those 65+ as an alternative would be of benefit in Switzerland, however, is not completely clear. In a recent study, between 2017 and 2019, 31% of IPD cases among those 65+ in Switzerland were caused by serotypes present in PCV13 (serotypes 8, 9N, and 22F, which are present in PPSV23, accounted for much of the remaining disease) [24]. This is in line with European data were 29% of IPD cases in those 65+ were caused by PCV13 serotypes (73% were caused by serotypes present in PPSV23) [6]. In a large, randomized, placebo-controlled Community-Acquired Pneumonia Immunization Trial in Adults (CAPiTA), PCV13 efficacy against all-type IPD in adults 65+ was 52% (75% for vaccine serotypes) and 45% against Community Acquired Pneumonia [44]. Considering, in a very simple way, the current situation in Switzerland with an average of 530 IPD cases per year among those 65+ [18]; if 31% of cases are caused by vaccine serotype strains and 75% could be prevented by PCV13 vaccination, this would mean that approximately 120 IPD cases per year could be prevented. There remains a need to better understand the effectiveness of PCV13 in protection against pneumococcal disease in the elderly, as well as the cost effectiveness of vaccination, and to make appropriate, evidence-based vaccination recommendations for this group.

### 4.4 Vaccine Perception and Intention to Vaccinate

Among unvaccinated individuals we found that reasons for not receiving pneumococcal vaccination were comparable between individuals reporting a health risk and those not reporting such a risk. The most common reasons for abstaining were “not enough information about the topic” and “not suggested by a doctor/healthcare provider”. At-risk respondents, however, were significantly less likely to report “not at increased risk due to chronic health conditions or age” as a reason for not being vaccinated compared to those not-at-risk (29% versus 4%), indicating that individuals with chronic health conditions are aware of their increased risk status. Furthermore, when unvaccinated individuals were asked about their intention to be vaccinated, significantly more at-risk individuals reported willingness to be vaccinated compared to those not-at-risk. A study of adults in the United States newly diagnosed with a chronic condition for which pneumococcal vaccination was indicated estimated that only 8% of individuals were vaccinated in the first year of follow-up and that this increased to only 20% after 5 years [45]. Similarly, a German study of adults with “high-risk” conditions found only ∼7-13% of patients were vaccinated for pneumococcus within 3 years of diagnosis [38]. Together, these findings suggest that interventions by a healthcare provider [46; 47; 48], specifically among at-risk individuals, may be an effective way to promote pneumococcal vaccination.

### 4.5 Study Limitations

An important limitation of this study is that the use of a cross-sectional design allows only correlations, but no causal effects to be determined. A nationwide vaccination registry would be ideal for rapid and accurate assessment of vaccination uptake. Unfortunately, such a representative registry is not yet available in Switzerland. Consequently, personal vaccination records are currently used to collect and evaluate vaccination data. While nearly 30% of contacted individuals submitted a questionnaire, only 16% submitted both a questionnaire and a vaccination record. While this is relatively low, it is not unusual for a study design based on response by mail and is comparable to similar studies which we have previously conducted [19; 20]. Importantly, however, the overall sample size was large enough to estimate pneumococcal vaccination coverage according to distinct age and risk strata of interest, as per study design.

With such a study design also comes the risk that individuals interested in a topic are more likely to participate. It is possible that individuals who have been vaccinated for pneumococcus or those having some previous experience with pneumococcal disease were more likely to participate. In this case, vaccination coverage would be overestimated, as would the frequency of individuals reporting some previous knowledge of pneumococcal disease. Furthermore, the study was conducted during the Covid-19 pandemic, with the first mailing sent just as the first cases were detected in Switzerland toward the end of February 2020 and the enrollment period beginning in March 2020, which was the beginning of the first lockdown period. Individuals may have been more likely to participate if they felt that they were at increased risk for severe respiratory disease.

Potential biases leading to underestimation of vaccination coverage may have occurred if vaccinations were not recorded after administration or if an individual was unable to provide a complete vaccination record. While we observed the highest level of vaccinations in 2020, our data are not complete for this year as the enrollment period was from March through December, and, for example, some early participants may have responded in March, but received a vaccination in July. This would lead to an underestimation of vaccination coverage in this year. Furthermore, self-reporting of health risks may have resulted in underreporting of, or inaccurately reporting a comorbidity. We additionally did not ask in our questionnaire about asplenia, sickle cell disease, lymphoma, leukemia, or myeloma, or cerebrospinal fluid leak as additional risk factors, which are indications for pneumococcal vaccination according to the Swiss FOPH.

### 4.6 Conclusions

Importantly, even considering potential limitations, our findings indicate that at-risk individuals in Switzerland are substantially undervaccinated against pneumococcus. Furthermore, our questionnaire data suggest that these individuals are aware of their increased risk, but that they do not have enough information on the topic to seek vaccination or have not been recommended a vaccination from their physician. Although coverage is increased by the presence of a health risk factor, in the best case less than 30% of adults with a given risk factor (immunosuppression) had received pneumococcal vaccination. Additionally, while age has been found to be an independent risk factor for pneumococcal pneumonia and rates of IPD in this age group are more than 3 times those of younger adults, vaccination rates among this group remain under 10%. These data support the need for effective programs to promote pneumococcal vaccination among those persons under 65 with chronic illnesses, and further indicate that an evaluation of the effectiveness of PCV13 vaccination in protecting those 65+ from IPD may be warranted.

## Data Availability

The dataset analyzed for this study is available upon request, without undue reservation, by contacting the study authors.

## 6 Author Contributions

KDZ and PL conceived and designed the study. KDZ and PL acquired study funding. KDZ and VB organized the database and analyzed the data, KDZ wrote the first draft of the manuscript. KDZ, VB, JSF, and PL critically revised the manuscript and approved the submitted version.

## 8 Acknowledgments

We would like to thank Anna Fraefel, Silvestro Superti-Furga, and Carlotta Superti-Furga for their help with data collection and organization during the study.

## References

[1] J.J.C. Drijkoningen, and G.G.U. Rohde, Pneumococcal infection in adults: burden of disease. Clinical Microbiology and Infection 20 (2014) 45–51.

[2] F. Blasi, M. Mantero, P. Santus, and P. Tarsia, Understanding the burden of pneumococcal disease in adults. Clinical Microbiology and Infection 18 (2012) 7–14.

[3] K.L. O’Brien, L.J. Wolfson, J.P. Watt, E. Henkle, M. Deloria-Knoll, N. McCall, E. Lee, K. Mulholland, O.S. Levine, and T. Cherian, Burden of disease caused by Streptococcus pneumoniae in children younger than 5 years: global estimates. The Lancet 374 (2009) 893–902.

[4] M.H. Kyaw, C.E. Rose, Jr., A.M. Fry, J.A. Singleton, Z. Moore, E.R. Zell, and C.G. Whitney, The influence of chronic illnesses on the incidence of invasive pneumococcal disease in adults. J Infect Dis 192 (2005) 377–86.

[5] A.J. van Hoek, N. Andrews, P.A. Waight, J. Stowe, P. Gates, R. George, and E. Miller, The effect of underlying clinical conditions on the risk of developing invasive pneumococcal disease in England. J Infect 65 (2012) 17–24.

[6] E.C.f.D.P.a. Control, Invasive pneumococcal disease - Annual Epidemiological Report for 2018. in: E.C.f.D.P.a. Control, (Ed.), Stockholm, 2018.

[7] Pneumokokkenerkrankungen 2012 [Pneumococcal Disease 2012]. in: S.F.O.o.P. Health, (Ed.), Swiss Federal Office of Public Health, 2014.

[8] Pneumokokken-Erkrankungen [Pneumococcal Illnesses]. in: S.F.O.f.P.H. (FOPH), (Ed.), Krankheiten A-Z [Diseases A-Z], Swiss Federal Office for Public Health, Bern, Switzerland, 2021.

[9] Pneumococcal vaccines WHO position paper - 2012 - recommendations. Vaccine 30 (2012) 4717–8.

[10] Empfohlene Impfungen für Personen mit einem erhöhten Risiko von Komplikationen oder invasiven Infektionen [Recommended Vaccinations for Persons with Increased Risk of Complications or Invasive Infections]. in: S.F.O.o.P. Health, (Ed.), Swiss Federal Office of Public Health, Bern, Switzerland, 2020.

[11] Schweizerischer Impfplan 2021 [Swiss Immunization Schedule 2021]. in: S.F.O.o.P. Health, (Ed.), Swiss Federal Office of Public Health, Federal Commission for Immunization Questions, Bern, Switzerland, 2021.

[12] Pneumokokkenimpfung: Empfehlungen zur Verhinderung von invasiven Pneumokokkenerkrankungen bei Risikogruppen [Pneumococcal Immunization: Recommendations to Reduce Invasive Pneumococcal Disease]. in: S.F.O.o.P. Health, (Ed.), Swiss Federal Office of Public Health, 2014.

[13] Pneumococcal Disease: Recommended vaccinations. in: E.C.f.D.P.a. Control, (Ed.), Vaccine Scheduler, European Centre for Disease Prevention and Control, 2021.

[14] Pneumococcal Vaccination: What Everyone Should Know. in: C.f.D.C.a. Prevention, (Ed.), Vaccines and Preventable Diseases, Centers for Disease Control and Prevention, 2020.

[15] Who should have the pneumococcal vaccine? in: N.H. Service, (Ed.), Vaccinations, National Health Service, 2019.

[16] E.C.f.D.P.a. Control, Pneumococcal Disease: Recommended vaccinations, Vaccine Scheduler.

[17] P. Castiglia, Recommendations for pneumococcal immunization outside routine childhood immunization programs in Western Europe. Adv Ther 31 (2014) 1011–1044.

[18] Pneumokokken: invasive Erkrankung [Pneumococcus: Invasive Disease]. in: S.F.O.o.P. Health, (Ed.), Zahlen zu Infektionskrankheiten [Numbers for Infectious Illnesses], Swiss Federal Office of Public Health, Bern, Switzerland, 2021.

[19] Kantonales Durchimpfungsmonitoring Schweiz [Cantonal Vaccination Monitoring, Switzerland]. in: S.F.O.o.P. Health, (Ed.), Swiss Federal Office of Public Health, Bern, Switzerland, 2021.

[20] V. Baroutsou, K.D. Zens, P. Sinniger, J. Fehr, and P. Lang, Analysis of Tick-borne Encephalitis vaccination coverage and compliance in adults in Switzerland, 2018. Vaccine 38 (2020) 7825–7833.

[21] S.F.S. Office, Regional Statistics, Federal Statistical Office Service ThemaKart, Neuchatel, Switzerland, 2020.

[22] S.F.S. Office, Demographic balance by age and canton, Swiss Federal Statistical Office Section Demography and Migration, Neuchatel, Switzerland, 2019.

[23] Schweizerischer Impfplan 2013 [Swiss Immunization Schedule 2013]. in: S.F.O.o.P. Health, (Ed.), Swiss Federal Office of Public Health, Federal Commission for Immunization Questions, Bern, Switzerland, 2013.

[24] O.R.-A. Oyewole, P. Lang, W.C. Albrich, K. Wissel, S.L. Leib, C. Casanova, and M. Hilty, The Impact of Pneumococcal Conjugate Vaccine (PCV) Coverage Heterogeneities on the Changing Epidemiology of Invasive Pneumococcal Disease in Switzerland, 2005–2019. Microorganisms 9 (2021) 1078.

[25] D.S. Fedson, L. Nicolas-Spony, P. Klemets, M. van der Linden, A. Marques, L. Salleras, and S.I. Samson, Pneumococcal polysaccharide vaccination for adults: new perspectives for Europe. Expert Review of Vaccines 10 (2011) 1143–1167.

[26] A. Kopp, O. Mangin, L. Gantzer, B. Lekens, G. Simoneau, M. Ravelomanantsoa, J. Evans, J.-F. Bergmann, and P. Sellier, Pneumococcal vaccination coverage in France by general practitioners in adults with a high risk of pneumococcal disease. Human Vaccines & Immunotherapeutics 17 (2021) 162–169.

[27] G. Gavazzi, B. Wazieres, B. Lejeune, and M. Rothan-Tondeur, Influenza and pneumococcal vaccine coverages in geriatric health care settings in france. Gerontology 53 (2007) 382–7.

[28] E. Rouveix, D. Gherissi Cherni, C. Dupont, A. Beauchet, H. Sordet Guepet, G. Gavazzi, and J. Gaillat, Streptococcus pneumoniae vaccinal coverage in hospitalized elderly patients in France. Med Mal Infect 43 (2013) 22–7.

[29] A.S. Delelis-Fanien, F. Séité, M. Priner, and M. Paccalin, [Vaccine coverage against influenza and pneumococcal infections in patients aged 65 and over: a survey on 299 outpatients]. Rev Med Interne 30 (2009) 656–60.

[30] U. Theidel, A. Kuhlmann, and A. Braem, Pneumococcal vaccination rates in adults in Germany: an analysis of statutory health insurance data on more than 850,000 individuals. Dtsch Arztebl Int 110 (2013) 743–50.

[31] D. Martinelli, S. Tafuri, G. Caputi, F. Fortunato, P. Reggio, C. Germinario, and R. Prato, Eight years of active proposal of pneumococcal 23-valent polysaccharide vaccine: Survey on coverage rate among elderly and chronic patients. American Journal of Infection Control 38 (2010) e8–e15.

[32] A.C. O’Halloran, P.J. Lu, and T. Pilishvili, Pneumococcal vaccination coverage among persons ≥65 years--United States, 2013. Vaccine 33 (2015) 5503–5506.

[33] Pneumococcal Polysaccharide Vaccine (PPV) coverage report, England, April 2017 to March 2018. in: P.H. England, (Ed.), Public Health England, 2018.

[34] W.C. Albrich, F. Rassouli, F. Waldeck, C. Berger, and F. Baty, Influence of Older Age and Other Risk Factors on Pneumonia Hospitalization in Switzerland in the Pneumococcal Vaccine Era. Frontiers in Medicine 6 (2019).

[35] K.M. Shea, J. Edelsberg, D. Weycker, R.A. Farkouh, D.R. Strutton, and S.I. Pelton, Rates of Pneumococcal Disease in Adults With Chronic Medical Conditions. Open Forum Infectious Diseases 1 (2014).

[36] B. Wyplosz, J. Fernandes, G. Goussiaume, J. Moïsi, J. Lortet-Tieulent, A. Vainchtock, C. Leboucher, and F. Raguideau, Adults at risk of pneumococcal disease in France. Infectious Diseases Now 51 (2021) 661–666.

[37] F.H.C.f.P. Health, Infections à pneumocoque : recommandations vaccinales pour les adultes [Pneumococcal infections: vaccine recommendations for adults], 2017.

[38] N. Schmedt, J. Schiffner-Rohe, R. Sprenger, J. Walker, C. von Eiff, and D. Häckl, Pneumococcal vaccination rates in immunocompromised patients—A cohort study based on claims data from more than 200,000 patients in Germany. PLOS ONE 14 (2019) e0220848.

[39] Influenza and pneumococcal vaccination coverage among persons aged > or =65 years and persons aged 18-64 years with diabetes or asthma--United States, 2003. MMWR Morb Mortal Wkly Rep 53 (2004) 1007–12.

[40] W.W. Williams, P.-J. Lu, A. O’Halloran, C.B. Bridges, D.K. Kim, T. Pilishvili, C.M. Hales, L.E. Markowitz, C. Centers for Disease, and Prevention, Vaccination coverage among adults, excluding influenza vaccination - United States, 2013. MMWR Morb Mortal Wkly Rep 64 (2015) 95–102.

[41] K. Grabowska, L. Högberg, P. Penttinen, Å. Svensson, and K. Ekdahl, Occurrence of invasive pneumococcal disease and number of excess cases due to influenza. BMC Infectious Diseases 6 (2006) 58.

[42] D.E. Morris, D.W. Cleary, and S.C. Clarke, Secondary Bacterial Infections Associated with Influenza Pandemics. Frontiers in Microbiology 8 (2017).

[43] Empfehlungen zur Pneumokokkenimpfung bei Kindern unter 5 Jahren: Wechsel vom 7-zum 13-valenten konjugierten Impfstoff [Recommendation for Pneumococcal Vaccination of Children Under 5: Change from 7-to 13-valent Conjugate Vaccine]. in: S.F.O.o.P. Health, (Ed.), Swiss Federal Office of Public Health, Bern, Switzerland, 2010.

[44] M.J.M. Bonten, S.M. Huijts, M. Bolkenbaas, C. Webber, S. Patterson, S. Gault, C.H. van Werkhoven, A.M.M. van Deursen, E.A.M. Sanders, T.J.M. Verheij, M. Patton, A. McDonough, A. Moradoghli-Haftvani, H. Smith, T. Mellelieu, M.W. Pride, G. Crowther, B. Schmoele-Thoma, D.A. Scott, K.U. Jansen, R. Lobatto, B. Oosterman, N. Visser, E. Caspers, A. Smorenburg, E.A. Emini, W.C. Gruber, and D.E. Grobbee, Polysaccharide Conjugate Vaccine against Pneumococcal Pneumonia in Adults. New England Journal of Medicine 372 (2015) 1114–1125.

[45] T. Petigara, and D. Zhang, Pneumococcal Vaccine Coverage in Adults Aged 19-64 Years, Newly Diagnosed With Chronic Conditions in the U.S. Am J Prev Med 54 (2018) 630–636.

[46] R. Eilers, P.F. Krabbe, and H.E. de Melker, Factors affecting the uptake of vaccination by the elderly in Western society. Prev Med 69 (2014) 224–34.

[47] M. Higuchi, K. Narumoto, T. Goto, and M. Inoue, Correlation between family physician’s direct advice and pneumococcal vaccination intention and behavior among the elderly in Japan: a cross-sectional study. BMC Family Practice 19 (2018) 153.

[48] J.M. Nagata, I. Hernández-Ramos, A.S. Kurup, D. Albrecht, C. Vivas-Torrealba, and C. Franco-Paredes, Social determinants of health and seasonal influenza vaccination in adults_J≥65 years: a systematic review of qualitative and quantitative data. BMC Public Health 13 (2013) 388.

