## Supplementary figures and images for "Pneumococcal Vaccination Coverage and Uptake Among Adults in Switzerland: A Nationwide Cross-Sectional Study of Vaccination Records"

### Supplementary Figures

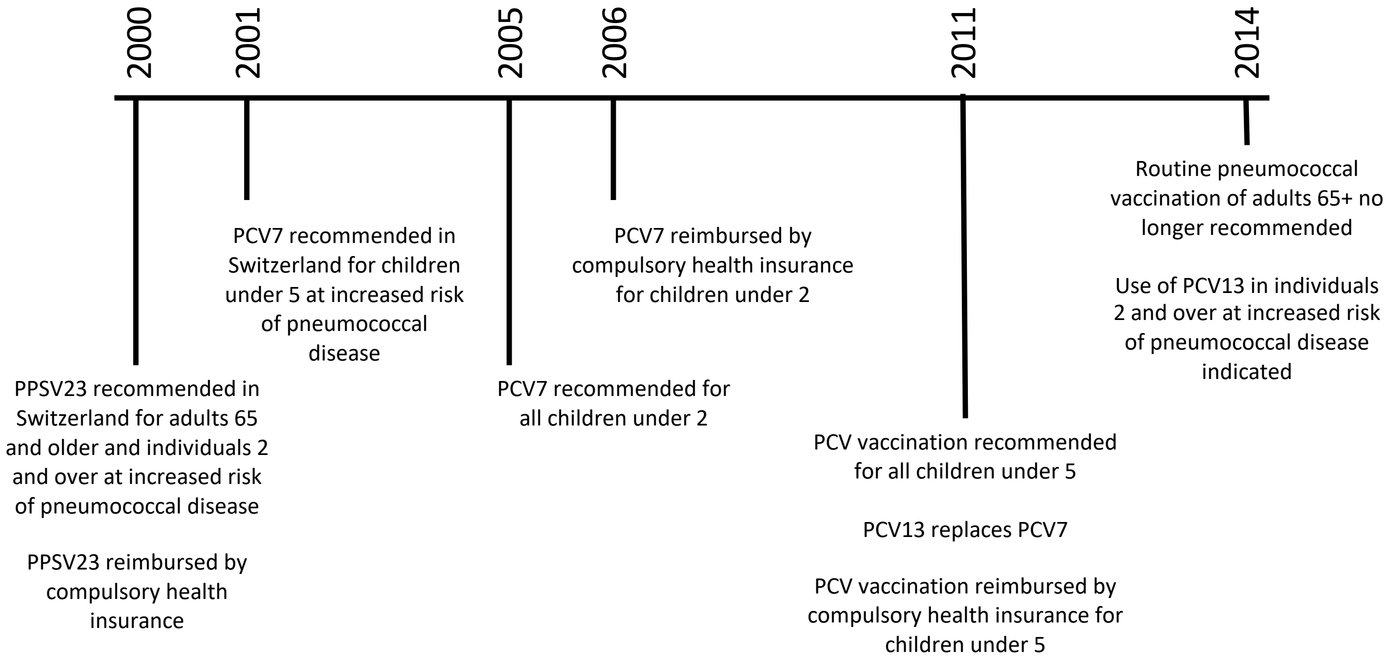

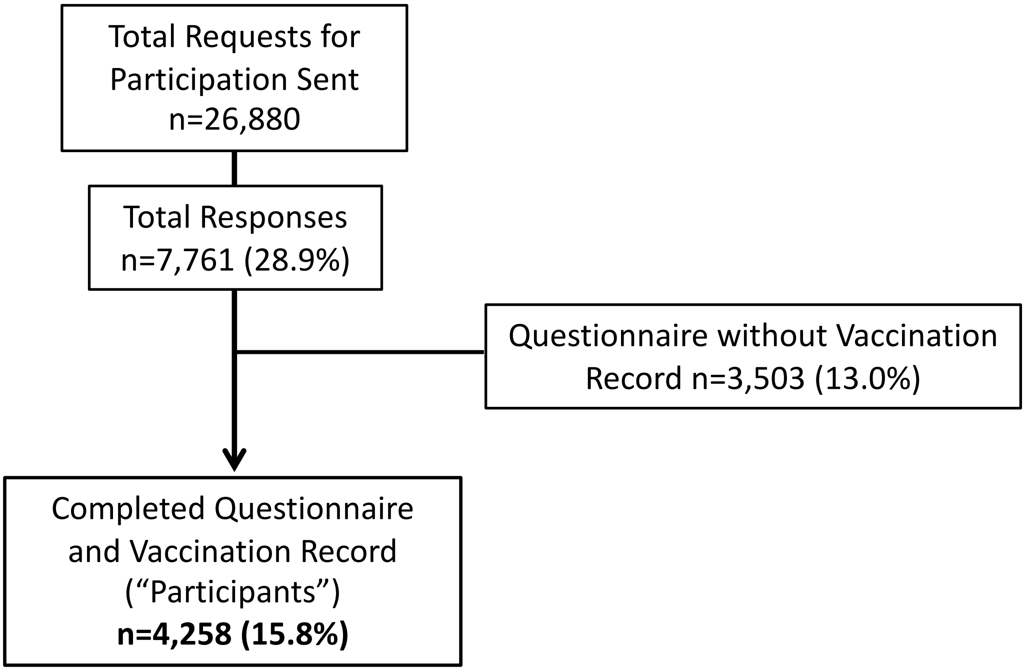

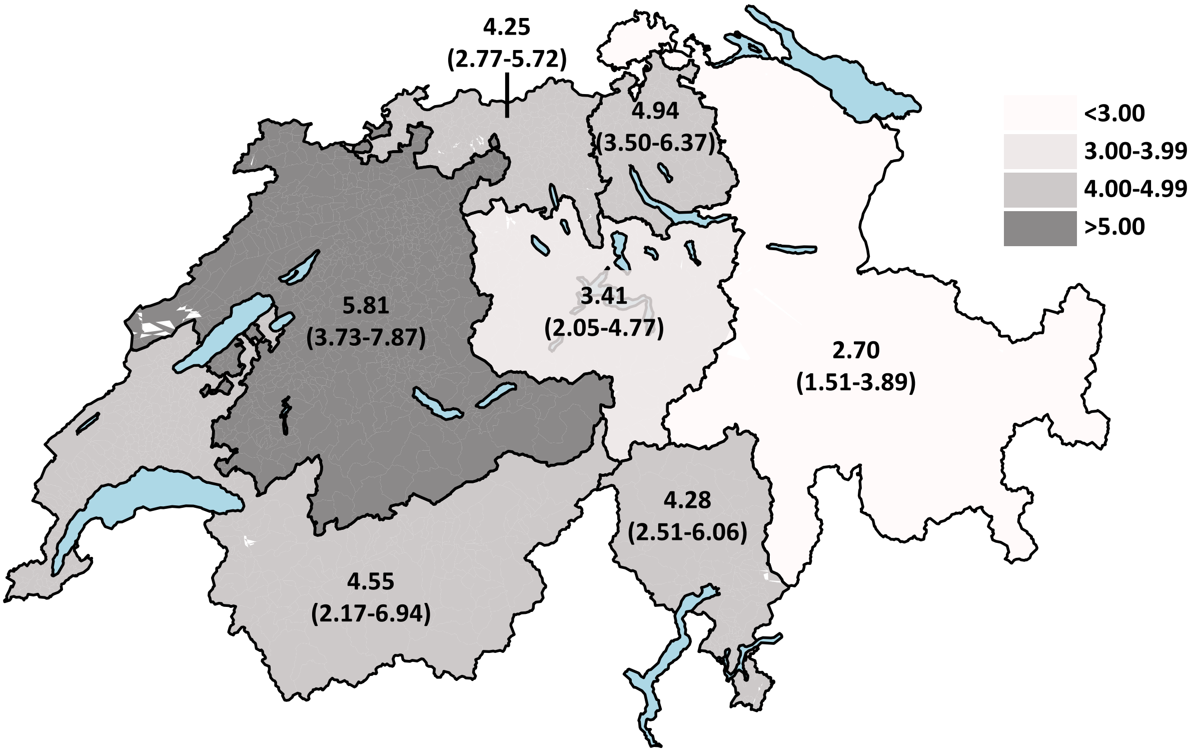
